## Supplementary figures and images for "The MFN2 Q367H variant reveals a novel pathomechanism connected to mtDNA-mediated inflammation"

### Supplemental Information

#### Supplemental Figure 1

A.

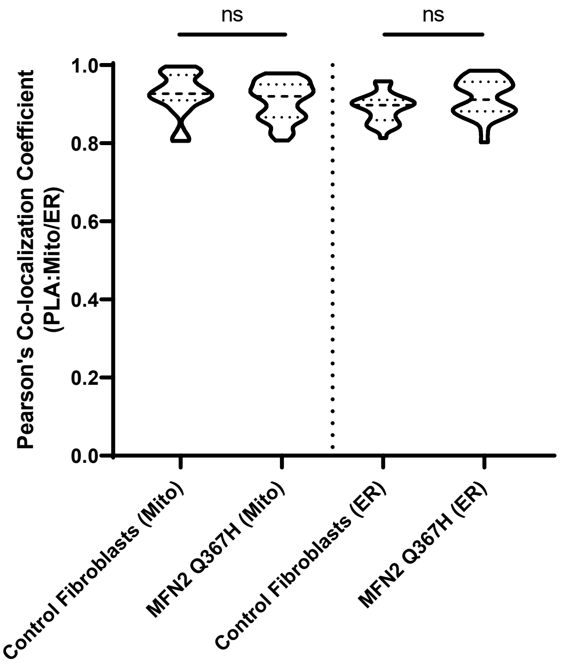

#### Supplemental Figure 2

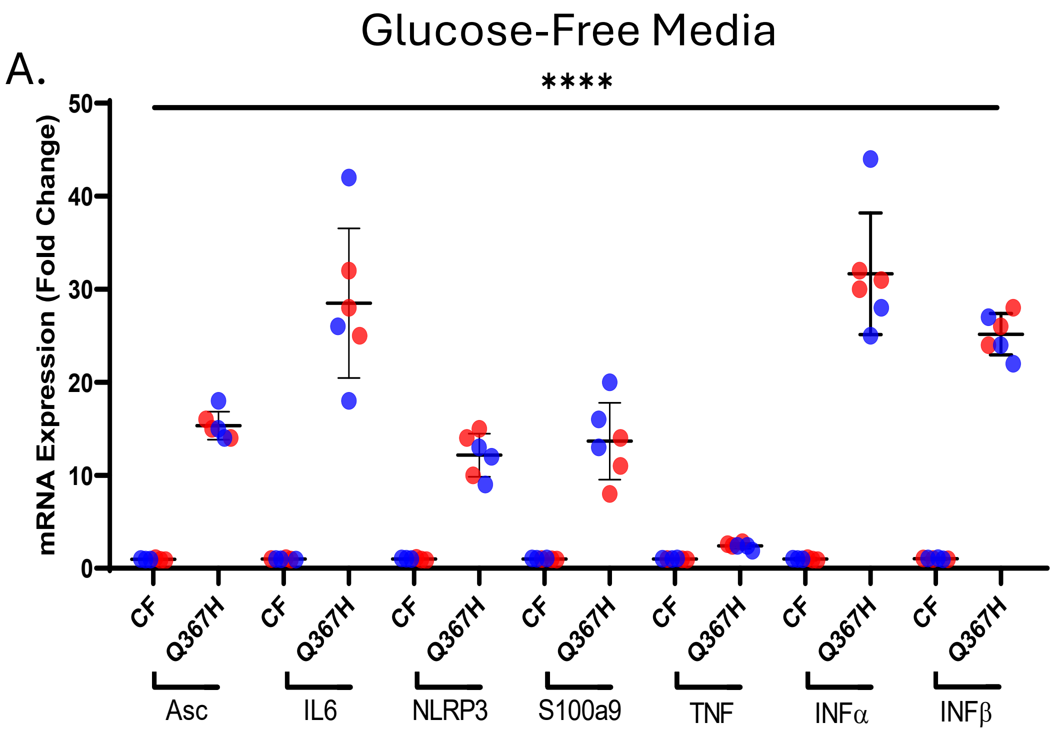

#### Supplemental Figure 3

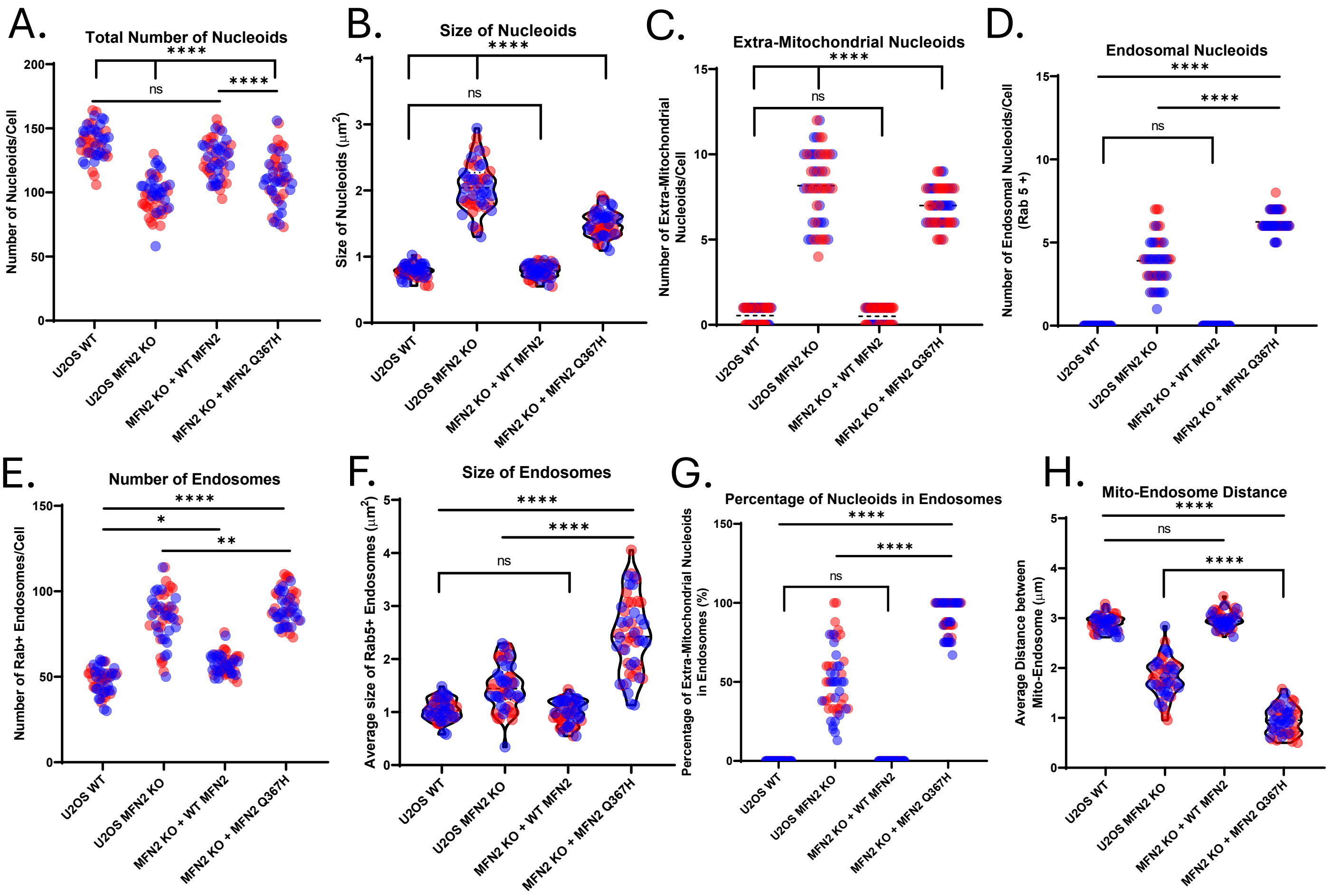
